# Childhood Sexual Abuse and Long-Term Risk of Self-Harm, Overdose, and Cardiovascular Disease

**DOI:** 10.64898/2026.07.09.26357627

**Authors:** Oluwasegun Akinyemi, Olububechukwu Eze, Mojisola Fasokun, Isaac Olaosebikan, Temitayo Ogundipe, Delia Singleton, Ayomide Ogunsakin, Samar Khalil, Kaelyn Gordon, Miriam Micheal, Kakra Hughes, Temitope Ogundare

**Author notes:** Corresponding Author: Oluwasegun Akinyemi MD PhD., 2041 Georgia Ave NW #4002, Washington, DC 20060, USA, Childhood Sexual Abuse and Long-Term Adult Health Risks.

## Abstract

**Importance:** Childhood sexual abuse (CSA) is linked to adverse psychiatric outcomes in adulthood, but evidence on its association with cardiovascular disease and mortality from large, diagnostically ascertained cohorts remains limited.

**Objective:** To assess the 10-year risk of all-cause mortality, suicide or self-harm, drug overdose or poisoning, and cardiovascular disease among patients with a diagnosed history of CSA compared with a matched unexposed cohort.

**Methods:** In this retrospective cohort study, we used deidentified electronic health record data from 68 health care organizations in the TriNetX US Collaborative Network. Patients diagnosed with confirmed or suspected childhood sexual abuse (CSA) before age 18 between January 1, 2003, and December 31, 2015, who had a subsequent adult encounter, were propensity score matched 1:1 with unexposed patients on age, sex, race and ethnicity, and baseline psychiatric and medical comorbidities (n = 9,083 per cohort). Outcomes—all-cause mortality, suicide or self-harm, drug overdose or poisoning, and cardiovascular disease—were assessed over 10 years from the index adult encounter using risk and time-to-event analyses to estimate risks, risk ratios, and hazard ratios.

**Results:** Among 18,166 matched patients (mean [SD] age, 19.0 [2.0] years; 14,813 [81.6%] female), CSA was associated with significantly elevated risk of suicide or self-harm (5.1% vs 2.8%; risk ratio [RR], 1.84; 95% CI, 1.57-2.16), drug overdose or poisoning (5.5% vs 3.7%; RR, 1.47; 95% CI, 1.28-1.69), and cardiovascular disease (12.3% vs 9.3%; RR, 1.31; 95% CI, 1.20-1.44), with concordant hazard ratios (all P < .001). All-cause mortality was numerically higher but not statistically significant (0.5% vs 0.4%; RR, 1.16; 95% CI, 0.75-1.79; P = .51).

**Conclusions and Relevance:** A diagnostically confirmed history of CSA was associated with substantially elevated 10-year risk of self-harm, overdose, and cardiovascular disease, independent of baseline demographic and psychiatric comorbidity. These findings support integrated psychiatric and cardiovascular screening for adult survivors of CSA and trauma-informed care extending beyond mental health services alone.

## Introduction

Childhood sexual abuse (CSA) affects an estimated 1 in 4 girls and 1 in 13 boys in the United States before age 18 [1], making it one of the most prevalent and clinically consequential adverse childhood experiences (ACEs) in both pediatric and adult medicine [2–4]. Unlike many ACEs that are diffuse in timing or severity, CSA is a clinically identifiable exposure when documented with a well-established association with downstream psychiatric morbidity, including depression, posttraumatic stress disorder, and substance use disorder [5–8]. What remains less well defined, particularly in large, diagnostically confirmed populations followed prospectively through adulthood, is the magnitude of CSA’s association with the physical health and mortality outcomes that ultimately drive population-level morbidity and health care spending [9–12].

The ACE framework, anchored by the original Kaiser-CDC study, established that cumulative childhood adversity confers a graded risk of adult disease, including cardiovascular disease, substance use disorder, and premature mortality [13–16]. However, the foundational ACE literature relies predominantly on retrospective self-report collected decades after exposure, introducing recall bias, survivorship bias, and an inability to disaggregate the specific contribution of sexual abuse from co-occurring forms of maltreatment [17–20]. Studies that isolate CSA as a discrete exposure have more often relied on convenience samples, clinical or forensic populations, or single-institution cohorts, limiting generalizability and statistical power to detect associations with comparatively rare outcomes such as suicide or premature cardiovascular disease [10, 21, 22].

The mechanisms linking CSA to adverse adult health are increasingly well characterized. Early-life trauma is associated with dysregulation of the hypothalamic-pituitary-adrenal axis, chronic low-grade inflammation, and accelerated allostatic load, each of which plausibly mediates downstream cardiometabolic risk [23–26]. In parallel, CSA is a well-established independent risk factor for suicidal ideation, suicide attempt, and substance use disorder, with effect sizes in prior work often exceeding those observed for other ACE subtypes [27–31]. Whether these associations persist when CSA is identified through structured clinical diagnosis rather than retrospective self-report, and when comparator groups are balanced on baseline psychiatric and medical comorbidity rather than left unadjusted, remains comparatively understudied.

The increasing availability of large, federated electronic health record (EHR) networks offers an opportunity to address these gaps. Diagnostic coding for confirmed and suspected CSA, captured prospectively in the medical record rather than reconstructed retrospectively, allows for cohort identification at a scale and with a temporal precision that survey-based designs cannot replicate [32, 33]. Propensity score matching on baseline demographic and psychiatric characteristics further improves control of confounding in the association between CSA and subsequent outcomes by balancing comorbidities that frequently co-occur with abuse disclosure, such as anxiety, posttraumatic stress disorder, and substance use disorder at baseline [6, 34, 35]. Such networks also permit a multidomain outcome profile, spanning mortality, self-directed violence, substance-related harm, and cardiovascular disease, within a single, consistently ascertained cohort followed over a clinically meaningful interval, clarifying whether the well-documented psychiatric signal is accompanied by a parallel, and potentially under-recognized, cardiovascular signal [9, 12, 28, 29].

In this study, we used a multicenter federated electronic health record network spanning dozens of health care organizations to construct a propensity score-matched cohort of patients with a diagnosed history of CSA before age 18 and a comparison cohort of patients without such a diagnosis, each followed from an index adult encounter for up to 10 years. We assessed the risk of all-cause mortality, suicide or self-harm, drug overdose or poisoning, and cardiovascular disease using risk, time-to-event, and instance-based analyses, with cohorts balanced on baseline demographic characteristics and prevalent psychiatric and medical comorbidity. We hypothesized that, even after this balancing, patients with a documented history of CSA would demonstrate significantly elevated risk across both psychiatric and cardiovascular outcome domains, reflecting the durable physiologic and behavioral sequelae of early-life sexual trauma, while recognizing that all-cause mortality risk may be difficult to detect given the relatively young age and limited follow-up duration achievable in this population.

Characterizing the magnitude and breadth of these associations in a large, diagnostically ascertained cohort followed into adulthood is intended to extend the existing ACE literature beyond retrospective self-report, to clarify the specific contribution of sexual abuse relative to other adversity subtypes, and ultimately to inform clinical screening practices, trauma-informed care models, and the allocation of behavioral and cardiovascular health resources for adult survivors of childhood sexual abuse.

## Methods

### Study Design and Data Source

We conducted a retrospective, propensity score-matched cohort study using TriNetX, a global federated health research network that provides access to deidentified electronic health record (EHR) data, including diagnoses, procedures, medications, and demographic information, aggregated across participating health care organizations (HCOs) [36, 37]. This analysis was performed within the US Collaborative Network, comprising 68 HCOs. The TriNetX platform was queried, and data were accessed for research purposes, between June 1, 2026, and June 20, 2026. This study followed the Strengthening the Reporting of Observational Studies in Epidemiology (STROBE) reporting guideline for cohort studies [38].

### Ethical Approval

This study used the TriNetX US Collaborative Network, which provides investigators with access to only aggregated, deidentified patient counts and statistical outputs; individual patient records are not accessible. Because the research involved no identifiable human subjects data, the study was exempted from institutional review board approval, and the requirement for informed consent was waived, consistent with prior TriNetX-based publications. The authors did not have access to information that could identify individual participants during or after data collection.

### Study Population

We identified two cohorts using structured diagnosis codes recorded between January 1, 2003, and December 31, 2015. The exposed cohort (CSA_Exposed) comprised patients with at least 1 diagnosis, recorded at 18 years of age or younger, of confirmed child sexual abuse (International Statistical Classification of Diseases, Tenth Revision, Clinical Modification [ICD-10-CM] code T74.22), suspected child sexual abuse (T76.22), or personal history of physical and sexual abuse in childhood (Z62.810), followed by at least 1 subsequent encounter (outpatient or emergency department visit) occurring at 18 years of age or older [39, 40]. The unexposed cohort (CSA_Unexposed) comprised patients with a qualifying pediatric encounter (outpatient or emergency department visit at ≤18 years) during the same calendar period, followed by a subsequent adult encounter, who did not carry a CSA or related maltreatment diagnosis code (T74, T76, or Z62.81 series) at any point in their recorded history and were not coded as deceased with an ill-defined cause of mortality (R99) before 18 years of age. Patients in both cohorts whose qualifying index event occurred more than 20 years before the analysis date were excluded.

### Index Event and Outcome Ascertainment

For each cohort, the index date was defined as the date of the first qualifying adult encounter following the exposure-defining (or comparator-defining) pediatric event. Outcomes were assessed in the time window beginning 1 day after the index date and ending 3650 days (10 years) later. Four outcomes were prespecified: all-cause mortality (ascertained via the TriNetX deceased demographic indicator); suicide or self-harm (ICD-10-CM codes X71–X83, T14.91, R45.851); drug overdose or poisoning (T36–T50); and cardiovascular disease (I00–I99, diseases of the circulatory system). For each outcome, patients with a record of that outcome before the start of the time window were excluded from the corresponding analysis to restrict estimates to incident events.

### Propensity Score Matching

Cohorts were balanced using 1:1 propensity score matching on age at index, sex, race and ethnicity (White, Black or African American, Asian, Hispanic or Latino), and baseline diagnoses of anxiety disorders, phobic anxiety disorders, posttraumatic stress disorder, substance use disorder, intentional self-harm, unspecified body region injury, overweight or obesity, diabetes mellitus, and essential hypertension, using a greedy nearest-neighbor algorithm with a caliper width of 0.1 pooled standard deviations of the logit of the propensity score, implemented within the TriNetX platform. Covariate balance before and after matching was assessed using standardized mean differences, with values below 0.1 considered indicative of adequate balance.

### Statistical Analysis

For each outcome, we calculated risk (the proportion of patients in each cohort experiencing the outcome within the time window), risk difference, and risk ratio, with 95% CIs constructed using a z test for risk difference. Time-to-event analysis was performed using a Cox proportional hazards model to estimate hazard ratios; patients without the outcome were censored at the date of their last recorded fact in the database. For outcomes with multiple recorded instances, the mean, median, and standard deviation of instance counts were compared using a 2-sample t test. Two-sided P < .05 was considered statistically significant. All analyses were conducted using the TriNetX Advanced Analytics platform (TriNetX, LLC) in June 2026.

## Results

### Cohort Characteristics

Among 9128 patients with a diagnosed history of childhood sexual abuse (CSA) and 5,294,899 unexposed patients meeting cohort query criteria, 1:1 propensity score matching on age, sex, race and ethnicity, and baseline psychiatric and medical comorbidity yielded 9083 matched patients per cohort (18,166 total). Before matching, the CSA-exposed cohort differed substantially from the unexposed cohort across nearly all measured characteristics, including posttraumatic stress disorder (17.5% vs 0.6%; standardized mean difference [SMD], 0.617), other anxiety disorders (24.2% vs 6.2%; SMD, 0.511), and overweight or obesity (19.9% vs 5.1%; SMD, 0.454), reflecting the psychiatric and medical burden previously associated with childhood maltreatment. After matching, covariate balance was excellent, with SMDs of 0.012 or less for every characteristic and between-cohort P values of .41 or greater across all comparisons (Table 1). The matched cohort had a mean (SD) age at index of 19.0 (2.0) years and was predominantly female (14,813 of 18,166 [81.6%]), consistent with the demographic profile of clinically ascertained CSA reported in prior literature.

**Table 1.** Baseline Characteristics of Patients with and Without a Documented History of Childhood Sexual Abuse, Before and After Propensity Score Matching.

| Characteristic | Before Propensity Score Matching |  |  | After Propensity Score Matching |  |  |
| --- | --- | --- | --- | --- | --- | --- |
|  | CSA-Exposed<br>(n = 9128) | CSA-Unexposed<br>(n = 5,294,899) | SMD | CSA-Exposed<br>(n = 9083) | CSA-Unexposed<br>(n = 9083) | SMD |
| *Demographics* |  |  |  |  |  |  |
| Age at index, mean (SD), y | 19.0 (2.0) | 19.5 (2.7) | 0.243 | 19.0 (2.0) | 19.0 (2.0) | 0.005 |
| White | 4856 (53.5) | 3,096,941 (58.5) | 0.137 | 4856 (53.5) | 4855 (53.5) | <0.001 |
| Black or African American | 2726 (30.0) | 965,178 (18.2) | 0.264 | 2724 (30.0) | 2723 (30.0) | <0.001 |
| Asian | 70 (0.8) | 180,155 (3.4) | 0.190 | 70 (0.8) | 71 (0.8) | 0.001 |
| Hispanic or Latino | 1544 (17.0) | 639,388 (12.1) | 0.129 | 1543 (17.0) | 1543 (17.0) | <0.001 |
| Female | 7411 (81.6) | 2,798,428 (52.9) | 0.609 | 7409 (81.6) | 7401 (81.5) | 0.002 |
| Male | 1670 (18.4) | 2,338,752 (44.2) | 0.607 | 1670 (18.4) | 1678 (18.5) | 0.002 |
| *Diagnoses* |  |  |  |  |  |  |
| Other anxiety disorders (ICD-10-CM F41) | 2198 (24.2) | 328,540 (6.2) | 0.511 | 2196 (24.2) | 2205 (24.3) | 0.002 |
| Phobic anxiety disorders (F40) | 144 (1.6) | 27,723 (0.5) | 0.102 | 144 (1.6) | 145 (1.6) | 0.001 |
| Posttraumatic stress disorder (F43.1) | 1589 (17.5) | 29,793 (0.6) | 0.617 | 1587 (17.5) | 1591 (17.5) | 0.001 |
| Mental/behavioral disorders due to psychoactive substance use (F10-F19) | 1113 (12.3) | 104,153 (2.0) | 0.405 | 1111 (12.2) | 1123 (12.4) | 0.004 |
| Intentional self-harm (X71-X83) | 320 (3.5) | 11,825 (0.2) | 0.244 | 318 (3.5) | 298 (3.3) | 0.012 |
| Injury of unspecified body region (T14) | 986 (10.9) | 157,685 (3.0) | 0.310 | 985 (10.8) | 992 (10.9) | 0.002 |
| Overweight and obesity (E66) | 1806 (19.9) | 268,424 (5.1) | 0.454 | 1804 (19.9) | 1798 (19.8) | 0.002 |
| Diabetes mellitus (E08-E13) | 222 (2.4) | 57,295 (1.1) | 0.101 | 221 (2.4) | 218 (2.4) | 0.002 |
| Essential (primary) hypertension (I10) | 303 (3.3) | 63,432 (1.2) | 0.141 | 302 (3.3) | 290 (3.2) | 0.007 |

**Abbreviations:** CSA, childhood sexual abuse; ICD-10-CM, International Classification of Diseases, Tenth Revision, Clinical Modification; SD, standard deviation; SMD, standardized mean difference.
Data are presented as No. (%) of patients unless otherwise indicated. SMD <0.10 is generally considered to indicate adequate covariate balance between cohorts; values ≥0.10 before matching are shown in the Before Propensity Score Matching columns to illustrate baseline imbalance that matching was designed to correct.
Before matching, the CSA-exposed (n = 9128) and CSA-unexposed (n = 5,294,899) cohorts reflect all patients meeting cohort query criteria. Counts for the age variable (9085 and 5,142,649 patients, respectively) reflect the subset with a non-missing age value, as reported directly by the TriNetX platform; percentage-based characteristics use the full cohort denominators above.
After matching, all between-cohort comparisons yielded $P \geq .41$ and $SMD \leq 0.012$ for every characteristic shown, indicating adequate covariate balance was achieved across demographic and clinical domains.

### Primary Outcomes

Over the 10-year follow-up window, CSA was associated with significantly elevated risk across 3 of 4 prespecified outcomes (Table 2). Risk of suicide or self-harm was more than 80% higher among CSA-exposed than unexposed patients (5.13% vs 2.79%; risk ratio [RR], 1.84; 95% CI, 1.57-2.16; hazard ratio [HR], 1.92; 95% CI, 1.63-2.26; P < .001), the largest effect size observed among the 4 outcomes assessed. Drug overdose or poisoning was similarly elevated (5.46% vs 3.72%; RR, 1.47; 95% CI, 1.28-1.69; HR, 1.52; 95% CI, 1.32-1.76; P < .001), as was incident cardiovascular disease (12.28% vs 9.34%; RR, 1.31; 95% CI, 1.20-1.44; HR, 1.40; 95% CI, 1.28-1.54; P < .001). All-cause mortality was numerically higher among CSA-exposed patients but did not reach statistical significance (0.49% vs 0.42%; RR, 1.16; 95% CI, 0.75-1.79; HR, 1.16; 95% CI, 0.75-1.80; P = .51), a finding consistent with limited statistical power in a young cohort with comparatively short observed follow-up rather than a true absence of association.

**Table 2.** Ten-Year Risk of Mortality, Self-Harm, Overdose, and Cardiovascular Disease Among Propensity-Matched Patients with and Without a Documented History of Childhood Sexual Abuse.

| Outcome | CSA-Exposed<br>No. (%) | CSA-Unexposed<br>No. (%) | Risk Ratio<br>(95% CI) | Hazard Ratio<br>(95% CI) | P Value |
| --- | --- | --- | --- | --- | --- |
| All-cause mortality | 44 (0.49) | 38 (0.42) | 1.16 (0.75-1.79) | 1.16 (0.75-1.80) | .51 |
| Suicide or self-harm | 389 (5.13) | 225 (2.79) | **1.84 (1.57-2.16)** | **1.92 (1.63-2.26)** | **<.001** |
| Drug overdose or poisoning | 443 (5.46) | 314 (3.72) | **1.47 (1.28-1.69)** | **1.52 (1.32-1.76)** | **<.001** |
| Cardiovascular disease | 963 (12.28) | 753 (9.34) | **1.31 (1.20-1.44)** | **1.40 (1.28-1.54)** | **<.001** |

**Abbreviations:** CI, confidence interval; CSA, childhood sexual abuse.
Risk is the proportion of patients in each cohort experiencing the outcome within 10 years following the index adult encounter. Risk ratios and 95% CIs were calculated using a z test for risk difference; hazard ratios were estimated from a Cox proportional hazards model, with patients without the outcome censored at the date of their last recorded fact in the TriNetX database.
The association with all-cause mortality was directionally consistent with the other three outcomes but did not reach statistical significance, likely reflecting limited statistical power given the relatively young cohort (mean age at index, 19.0 years) and the comparatively short observed follow-up rather than a true absence of effect.

### Stratified Analyses

Sex-stratified analyses demonstrated directionally consistent elevations in risk for suicide or self-harm, drug overdose or poisoning, and cardiovascular disease among both female and male patients with CSA (Table 3). Point estimates were numerically larger among female patients for suicide or self-harm (RR, 2.11; 95% CI, 1.75-2.54) than among male patients (RR, 1.94; 95% CI, 1.32-2.85), whereas drug overdose or poisoning risk was numerically larger among male patients (RR, 2.18; 95% CI, 1.42-3.34) than female patients (RR, 1.59; 95% CI, 1.36-1.86). The cardiovascular disease risk ratio did not reach statistical significance in the smaller male stratum alone (RR, 1.24; 95% CI, 0.99-1.56), despite a statistically significant hazard ratio (HR, 1.43; 95% CI, 1.12-1.82), a pattern consistent with reduced power rather than a true sex difference in effect.

**Table 3.**
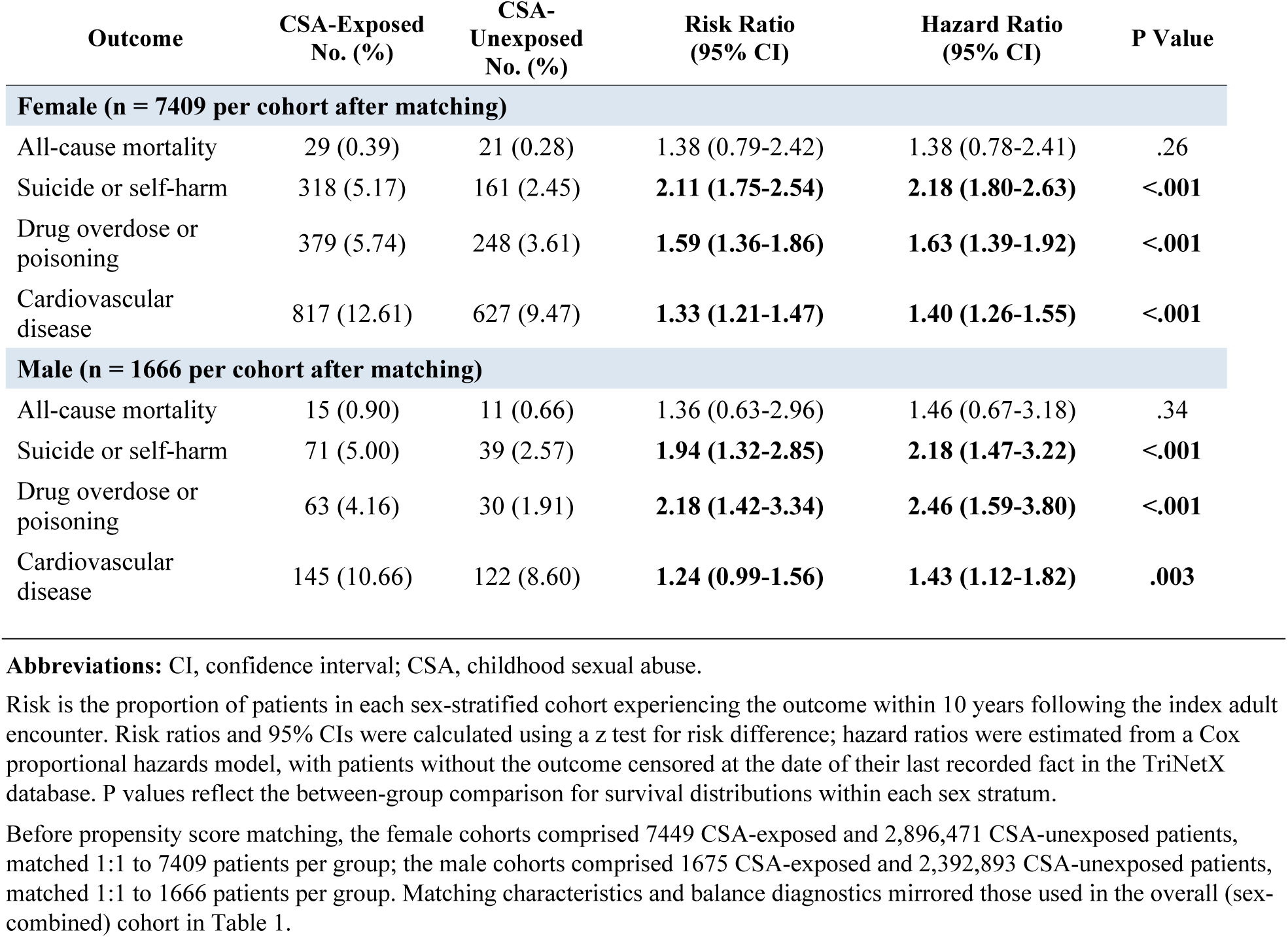
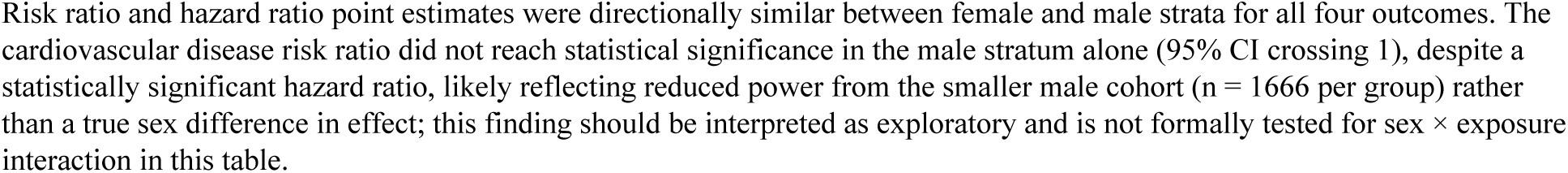
Ten-Year Risk of Mortality, Self-Harm, Overdose, and Cardiovascular Disease Among Patients with and Without a Documented History of Childhood Sexual Abuse, Stratified by Sex.

| Outcome | CSA-Exposed<br>No. (%) | CSA-Unexposed<br>No. (%) | Risk Ratio<br>(95% CI) | Hazard Ratio<br>(95% CI) | P Value |
| --- | --- | --- | --- | --- | --- |
| **Female (n = 7409 per cohort after matching)** |  |  |  |  |  |
| All-cause mortality | 29 (0.39) | 21 (0.28) | 1.38 (0.79-2.42) | 1.38 (0.78-2.41) | .26 |
| Suicide or self-harm | 318 (5.17) | 161 (2.45) | **2.11 (1.75-2.54)** | **2.18 (1.80-2.63)** | **<.001** |
| Drug overdose or poisoning | 379 (5.74) | 248 (3.61) | **1.59 (1.36-1.86)** | **1.63 (1.39-1.92)** | **<.001** |
| Cardiovascular disease | 817 (12.61) | 627 (9.47) | **1.33 (1.21-1.47)** | **1.40 (1.26-1.55)** | **<.001** |
| **Male (n = 1666 per cohort after matching)** |  |  |  |  |  |
| All-cause mortality | 15 (0.90) | 11 (0.66) | 1.36 (0.63-2.96) | 1.46 (0.67-3.18) | .34 |
| Suicide or self-harm | 71 (5.00) | 39 (2.57) | **1.94 (1.32-2.85)** | **2.18 (1.47-3.22)** | **<.001** |
| Drug overdose or poisoning | 63 (4.16) | 30 (1.91) | **2.18 (1.42-3.34)** | **2.46 (1.59-3.80)** | **<.001** |
| Cardiovascular disease | 145 (10.66) | 122 (8.60) | **1.24 (0.99-1.56)** | **1.43 (1.12-1.82)** | **.003** |

**Abbreviations:** CI, confidence interval; CSA, childhood sexual abuse.
Risk is the proportion of patients in each sex-stratified cohort experiencing the outcome within 10 years following the index adult encounter. Risk ratios and 95% CIs were calculated using a z test for risk difference; hazard ratios were estimated from a Cox proportional hazards model, with patients without the outcome censored at the date of their last recorded fact in the TriNetX database. P values reflect the between-group comparison for survival distributions within each sex stratum.
Risk ratio and hazard ratio point estimates were directionally similar between female and male strata for all four outcomes. The cardiovascular disease risk ratio did not reach statistical significance in the male stratum alone (95% CI crossing 1), despite a statistically significant hazard ratio, likely reflecting reduced power from the smaller male cohort (n = 1666 per group) rather than a true sex difference in effect; this finding should be interpreted as exploratory and is not formally tested for sex × exposure interaction in this table.

Race and ethnicity-stratified analyses likewise demonstrated consistent elevations in risk of suicide or self-harm and drug overdose or poisoning across White, Black or African American, and Hispanic or Latino patients (all P ≤ .004), with the largest point estimates for both outcomes observed among Hispanic or Latino patients (suicide or self-harm RR, 2.32; 95% CI, 1.47-3.66; drug overdose or poisoning RR, 2.08; 95% CI, 1.34-3.23) (Table 4). The cardiovascular disease association reached statistical significance among White patients (RR, 1.29; 95% CI, 1.14-1.45) and was supported by a significant hazard ratio among Black or African American patients (HR, 1.27; 95% CI, 1.08-1.50) despite a risk ratio that approached but did not cross the null (RR, 1.11; 95% CI, 0.95-1.29). The cardiovascular disease risk ratio did not reach significance in the smaller Hispanic or Latino stratum (RR, 1.24; 95% CI, 0.95-1.61). All-cause mortality did not reach statistical significance within any individual racial or ethnic stratum, and this outcome was not generated for the Hispanic or Latino stratum in the underlying analysis. These stratified findings were not formally tested for sex or race and ethnicity by exposure interaction and should be interpreted as exploratory, hypothesis-generating analyses rather than confirmatory subgroup effects.

**Table 4.**
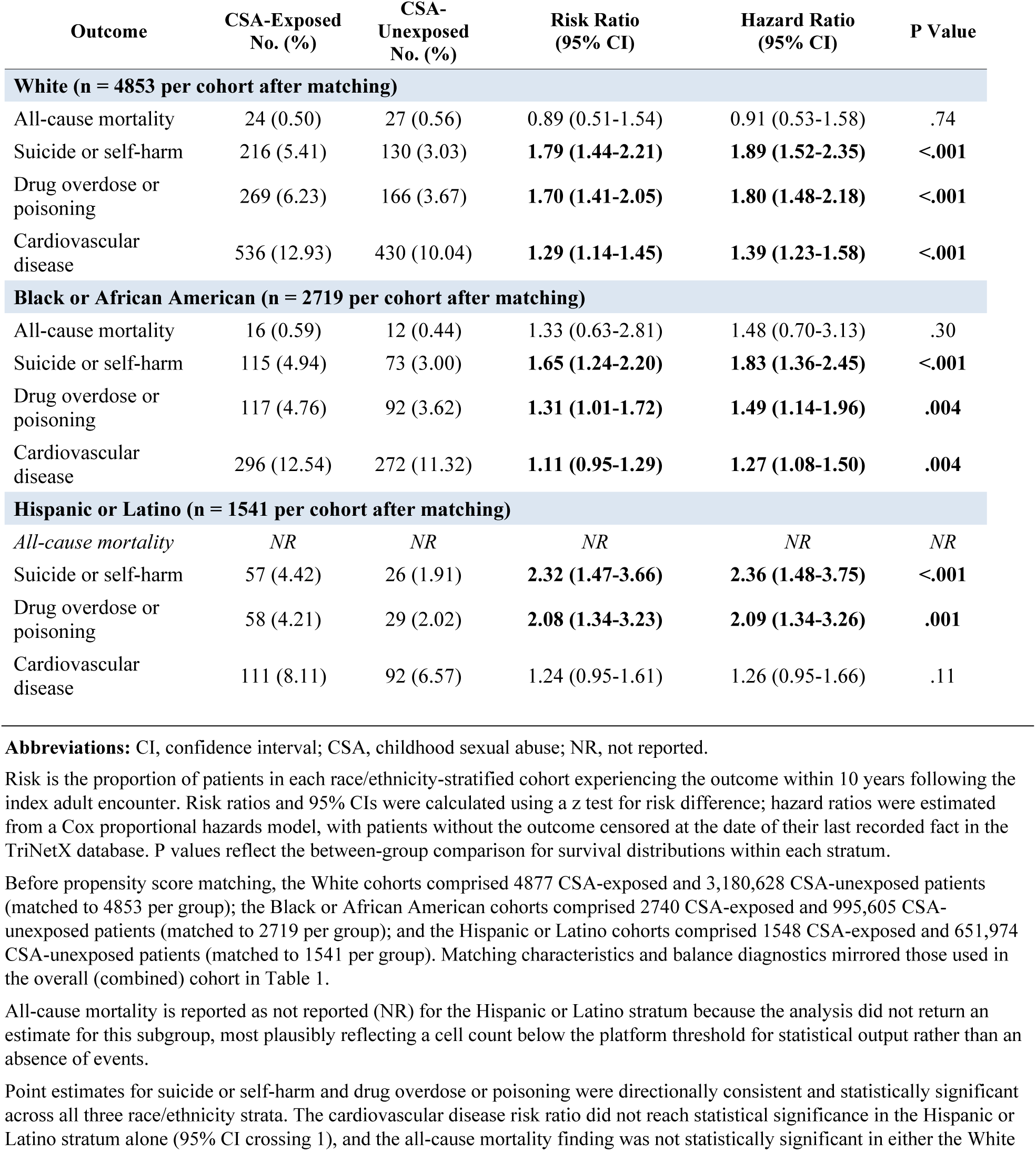
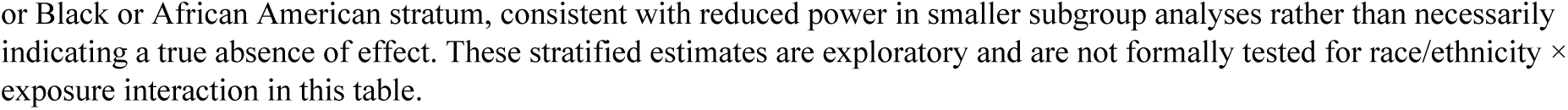
Ten-Year Risk of Mortality, Self-Harm, Overdose, and Cardiovascular Disease Among Patients with and Without a Documented History of Childhood Sexual Abuse, Stratified by Race and Ethnicity.

| Outcome | CSA-Exposed<br>No. (%) | CSA-Unexposed<br>No. (%) | Risk Ratio<br>(95% CI) | Hazard Ratio<br>(95% CI) | P Value |
| --- | --- | --- | --- | --- | --- |
| **White (n = 4853 per cohort after matching)** |  |  |  |  |  |
| All-cause mortality | 24 (0.50) | 27 (0.56) | 0.89 (0.51-1.54) | 0.91 (0.53-1.58) | .74 |
| Suicide or self-harm | 216 (5.41) | 130 (3.03) | **1.79 (1.44-2.21)** | **1.89 (1.52-2.35)** | **<.001** |
| Drug overdose or poisoning | 269 (6.23) | 166 (3.67) | **1.70 (1.41-2.05)** | **1.80 (1.48-2.18)** | **<.001** |
| Cardiovascular disease | 536 (12.93) | 430 (10.04) | **1.29 (1.14-1.45)** | **1.39 (1.23-1.58)** | **<.001** |
| **Black or African American (n = 2719 per cohort after matching)** |  |  |  |  |  |
| All-cause mortality | 16 (0.59) | 12 (0.44) | 1.33 (0.63-2.81) | 1.48 (0.70-3.13) | .30 |
| Suicide or self-harm | 115 (4.94) | 73 (3.00) | **1.65 (1.24-2.20)** | **1.83 (1.36-2.45)** | **<.001** |
| Drug overdose or poisoning | 117 (4.76) | 92 (3.62) | **1.31 (1.01-1.72)** | **1.49 (1.14-1.96)** | **.004** |
| Cardiovascular disease | 296 (12.54) | 272 (11.32) | **1.11 (0.95-1.29)** | **1.27 (1.08-1.50)** | **.004** |
| **Hispanic or Latino (n = 1541 per cohort after matching)** |  |  |  |  |  |
| All-cause mortality | NR | NR | NR | NR | NR |
| Suicide or self-harm | 57 (4.42) | 26 (1.91) | **2.32 (1.47-3.66)** | **2.36 (1.48-3.75)** | **<.001** |
| Drug overdose or poisoning | 58 (4.21) | 29 (2.02) | **2.08 (1.34-3.23)** | **2.09 (1.34-3.26)** | **.001** |
| Cardiovascular disease | 111 (8.11) | 92 (6.57) | 1.24 (0.95-1.61) | 1.26 (0.95-1.66) | .11 |

**Abbreviations:** CI, confidence interval; CSA, childhood sexual abuse; NR, not reported.
Risk is the proportion of patients in each race/ethnicity-stratified cohort experiencing the outcome within 10 years following the index adult encounter. Risk ratios and 95% CIs were calculated using a z test for risk difference; hazard ratios were estimated from a Cox proportional hazards model, with patients without the outcome censored at the date of their last recorded fact in the TriNetX database. P values reflect the between-group comparison for survival distributions within each stratum.
All-cause mortality is reported as not reported (NR) for the Hispanic or Latino stratum because the analysis did not return an estimate for this subgroup, most plausibly reflecting a cell count below the platform threshold for statistical output rather than an absence of events.
Point estimates for suicide or self-harm and drug overdose or poisoning were directionally consistent and statistically significant across all three race/ethnicity strata. The cardiovascular disease risk ratio did not reach statistical significance in the Hispanic or Latino stratum alone (95% CI crossing 1), and the all-cause mortality finding was not statistically significant in either the White
or Black or African American stratum, consistent with reduced power in smaller subgroup analyses rather than necessarily indicating a true absence of effect. These stratified estimates are exploratory and are not formally tested for race/ethnicity × exposure interaction in this table.

## Discussion

### Principal Findings

In this propensity score-matched cohort study of more than 18,000 patients drawn from a federated electronic health record network, a diagnosed history of childhood sexual abuse (CSA) was associated with significantly elevated 10-year risk of suicide or self-harm, drug overdose or poisoning, and cardiovascular disease, even after balancing cohorts on baseline demographic characteristics and prevalent psychiatric and medical comorbidity. The association with all-cause mortality was directionally consistent but did not reach statistical significance, likely reflecting limited statistical power given the young age and comparatively short observed follow-up of this cohort rather than a true absence of effect. Together, these findings extend the adverse childhood experience (ACE) literature by demonstrating that a diagnostically ascertained, code-confirmed CSA exposure carries a multidomain health burden spanning both psychiatric and cardiovascular outcomes, with the magnitude of the suicide and self-harm association in particular exceeding that typically reported for composite ACE exposure.

### Significance

The cardiovascular finding is notable. While the psychiatric sequelae of CSA are well established, prospective, diagnostically confirmed evidence linking CSA specifically to incident cardiovascular disease in young adulthood remains comparatively sparse [41, 42]. A risk ratio exceeding 1.3, observed within a decade of an index adult encounter in a cohort with a mean age of approximately 19 years, suggests that allostatic and inflammatory sequelae of early-life trauma may manifest in measurable cardiovascular morbidity earlier in the life course than has traditionally been appreciated, with implications for primary prevention counseling in trauma-informed primary care [26, 41, 43–45].

### Strengths

This study has several notable strengths. The use of a large, multicenter federated network spanning 68 health care organizations provided substantial statistical power and generalizability beyond single-institution or convenience samples. CSA exposure was ascertained through structured clinical diagnosis rather than retrospective self-report, reducing recall bias inherent to survey-based ACE research. Propensity score matching achieved excellent covariate balance, with standardized differences below 0.02 across all measured demographic and comorbidity variables, and outcomes were assessed using complementary risk, time-to-event, and instance-based analytic approaches.

### Limitations

This study also has important limitations. First, the comparison cohort was defined only by the absence of a CSA or related maltreatment diagnosis code and was not screened for other forms of childhood adversity; residual confounding by unmeasured trauma in the comparator group would be expected to bias estimates toward the null, suggesting the true associations may be larger than observed [46, 47]. Second, ascertainment bias is a central concern: patients coded for CSA were, by definition, already engaged with a health system in a manner that prompted disclosure or clinical suspicion, which may correlate with unmeasured factors influencing subsequent health care-seeking behavior and diagnostic capture of outcomes [48]. Third, diagnosis codes do not capture abuse severity, chronicity, perpetrator relationship, or corroboration, precluding dose-response analysis [49]. Fourth, EHR-based ascertainment is subject to differential loss to follow-up and informative censoring, as patients are censored at their last recorded encounter, which may differ systematically between cohorts [50–53]. Fifth, the relatively short median follow-up limits inference about lifetime mortality risk and later-onset cardiovascular disease. Sixth, granular socioeconomic and family-environment data, important confounders in trauma research, are not consistently captured in claims-based EHR networks [54–58]. Finally, multiple outcomes were assessed without formal correction for multiplicity; however, the three statistically significant primary associations each had P < .001 and would remain significant after Bonferroni correction for the four primary comparisons (adjusted threshold, P < .0125), and the magnitude and consistency of effect sizes across analytic approaches suggest these findings are unlikely to reflect chance alone [59].

### Future Directions

Future research should prioritize linkage of EHR-based cohorts to corroborated abuse severity data, extended follow-up into mid- and late-adulthood to clarify lifetime mortality risk, and dose-response analyses incorporating abuse chronicity and co-occurring maltreatment subtypes. Studies evaluating whether trauma-informed care interventions or early behavioral health engagement attenuate the cardiovascular and overdose risk identified here would directly inform clinical practice.

## Conclusions

A diagnostically confirmed history of childhood sexual abuse was associated with substantially elevated 10-year risk of self-harm, overdose, and cardiovascular disease in this large, propensity score-matched cohort, underscoring the need for integrated psychiatric and cardiovascular screening in adult survivors of childhood sexual abuse.

## Acknowledgment

Artificial intelligence assistance was provided by Claude (claude-sonnet-4-6; Anthropic, PBC, San Francisco, CA) for table formatting and editorial editing of the manuscript text. All intellectual content, data analysis, interpretations, and conclusions are solely those of the authors. The authors take full responsibility for the integrity of the AI-assisted content presented in this manuscript.

## Data Availability

The data underlying this study were obtained from the TriNetX US Collaborative Network under a data use agreement and are not publicly available owing to restrictions imposed by that agreement. TriNetX provides only aggregated, deidentified patient counts and statistical outputs, and the authors are not permitted to redistribute patient-level data. Qualified researchers may request access directly from TriNetX (https://trinetx.com); the cohort definitions, diagnosis codes, and analytic parameters required to reproduce these analyses are provided in full within the manuscript and its tables.

## Author Contributions

O.A. conceptualized and designed the study. O.E and O.A conducted the analyses, while O.A, M.F, I.O, T.O drafted the manuscript. All authors contributed to interpretation of the data, critically revised the manuscript for important intellectual content, and approved the final version for submission. O.E. and O.A had full access to all study data and takes responsibility for the integrity of the data and the accuracy of the analysis.

